# Blood RNA and protein biomarkers are associated with vaping and dual use, and prospective health outcomes

**DOI:** 10.1101/2022.09.19.22280093

**Authors:** Andrew Gregory, Zhonghui Xu, Katherine Pratte, Seth Berman, Robin Lu, Rahul Suryadevara, Robert Chase, Jeong H. Yun, Aabida Saferali, Craig P. Hersh, Edwin K. Silverman, Russell P. Bowler, Laura E. Crotty Alexander, Adel Boueiz, Peter J. Castaldi

## Abstract

**Background:** Electronic nicotine delivery systems (ENDS) are driving an epidemic of vaping. Identifying biomarkers of vaping and dual use (concurrent vaping and smoking) will facilitate studies of the health effects of vaping. To identify putative biomarkers of vaping and dual use, we performed association analysis in an observational cohort of 3,892 COPDGene study participants with blood transcriptomics and/or plasma proteomics data and self-reported current vaping and smoking behavior.

**Methods:** Biomarkers of vaping and dual use were identified through differential expression analysis and related to prospective health events over six years of follow-up. To assess the predictive accuracy of multi-biomarker panels, we constructed predictive models for vaping and smoking categories and prospective health outcomes.

**Results:** We identified three transcriptomic and three proteomic associations with vaping, and 90 transcriptomic and 100 proteomic associations to dual use. Many of these vaping or dual use biomarkers were significantly associated with prospective health outcomes, such as FEV1 decline (three transcripts and 62 proteins), overall mortality (18 transcripts and 73 proteins), respiratory mortality (two transcripts and 23 proteins), respiratory exacerbations (13 proteins) and incident cardiovascular disease (24 proteins). Multimarker models showed good performance discriminating between vaping and smoking behavior and produced informative, modestly powerful predictions of future FEV1 decline, mortality, and respiratory exacerbations.

**Conclusions:** In summary, vaping and dual use are associated with RNA and protein blood-based biomarkers that are also associated with adverse health outcomes.

## Introduction

The high prevalence of electronic cigarette use (vaping) among both adolescents[1] and adult cigarette users[2] is a major public health concern. Electronic cigarettes, also referred to as electronic nicotine delivery systems (ENDS), were initially marketed as a means of smoking reduction, with the hope that their effects on the health of cigarette users would be a net positive. However, vaping is associated with acute respiratory symptoms[3–5] and lung injury[6,7] and is likely to be associated with long-term adverse health effects. Vaping companies are targeting young people as a profitable demographic and promoting ENDS use beyond the population of established cigarette users, resulting in an ongoing epidemic of vaping in adolescents and non-cigarette user adults that is clearly detrimental to public health[8–10].

The Family Smoking Prevention and Tobacco Control Act of 2009 gave the Food and Drug Administration (FDA) regulatory oversight over tobacco products which, by the FDA Deeming Regulation of 2016, includes ENDS. Thus, the FDA is responsible for reviewing the safety of all ENDS products introduced to market after February 15, 2007. To facilitate this review process, it is important to determine whether there are validated biomarkers of ENDS use and combined use of ENDS and cigarettes (dual use). While biomarkers of vaping and dual use have been reported, few if any vaping biomarkers have been assessed for their association to prospective health outcomes, and the predictive performance of multi-biomarker panels has not been assessed. In this paper, the term biomarker refers to molecular measurements tested for association to vaping or smoking exposure, which we distinguish from validated biomarkers in which a specific association has been demonstrated in multiple studies. Validated biomarkers that are also predictive of prospective health events can be considered for use as surrogate outcomes in safety and health assessments[11,12] which would be a useful tool for evaluating the health effects of an increasingly diverse array of ENDS products. Thus, the goal of this study is to establish a set of candidate biomarkers for future validation and further study as potential surrogate outcomes.

We hypothesized that vaping and dual use are associated to multi-omic biomarkers, and that a subset of these biomarkers would be associated with the development of pulmonary and cardiovascular disease. We conducted high throughput transcriptomic and proteomic analyses that identified over 100 biomarkers associated with vaping and dual use in a cohort of adult current and former cigarette users in the COPDGene study. We tested these biomarkers for association with prospective spirometric changes, mortality, and incident cardiovascular disease, identifying dozens of significantly associated biomarkers. We also investigated the extent to which multimarker panels can predict vaping and smoking status and the development of prospective health outcomes.

## Methods

### Study description

The COPDGene study recruited 10,198 non-Hispanic White and Black cigarette users with at least 10 pack-years of lifetime cigarette smoking history at 21 U.S. clinical centers (NCT00608764, www.copdgene.org)[13] between 2007 and 2011. COPDGene obtained five-year follow-up and is currently obtaining 10-year follow-up of available subjects to collect longitudinal data on study participants. A total of 6,756 subjects completed their five-year study visit (Visit 2) and the 10-year study visit (Visit 3) is ongoing at time of writing. Data collected includes questionnaires, spirometry, and self-reported smoking and vaping behaviors. At Visit 2, whole-blood RNA sequencing (RNA-seq), plasma protein measurements (SOMAscan version 4.0 (5.0K) assay for human plasma), and plasma cotinine were obtained from a subset of subjects. Throughout the study, subjects were also regularly contacted via the phone or Internet as part of the COPDGene Longitudinal Follow-up (LFU) program to assess interval events, including respiratory exacerbations, incident comorbidities, and mortality. Definitions of respiratory exacerbations, incident comorbidities, and mortality assessment are included in the supplementary materials of the online preprint. Written informed consents were obtained for all subjects, and each clinical site obtained institutional review board approval.

### Vaping and smoking groups

Starting from Visit 2, vaping was determined by self-report including ever use, current use, intensity, cartridge size, brand, and flavoring. The four comparison groups for biomarker discovery (vapers, current cigarette users, former cigarette users, and dual users) were defined based on the subjects’ questionnaire responses at Visit 2. Vapers were subjects who reported using at least one e-cigarette within the prior week and had a history of smoking tobacco cigarettes, but not within the last 30 days. Current cigarette users reported current smoking with an average of at least one cigarette per day without any e-cigarette use. Dual users were vapers who also reported current smoking, and former cigarette users were defined as those who reported a history of smoking but did not meet criteria for current smoking or vaping. In most of the reported analyses, former cigarette users are used as the reference group. Differences in baseline characteristics between groups were assessed with Kruskal-Wallis and chi-square tests for continuous and categorical measurements, respectively.

### RNA-sequencing

The extraction protocol for total blood RNA was performed either manually or with the Qiagen QIAcube extraction robot according to the company’s standard operating procedure. RNA samples with an RNA integrity number (RIN) > 6 and a concentration of ≥ 25 μg/μl were sequenced. Library preparation was performed with the Illumina TruSeq Stranded Total RNA with Ribo-Zero Globin kit (Illumina, Inc., San Diego, CA). Samples were sequenced to an average depth of 20 million 75 base pairs (bp) paired end reads on Ilumina HiSeq 2500 sequencers. Samples were sequenced to an average depth of 20 million 75 base pairs (bp) paired end reads on Ilumina HiSeq 2500 sequencers. Reads were aligned to the GRCh38 genome using the STAR (version 2.5.2b) align. Gene annotations and transcript GTF were downloaded from the Biomart Ensembl database (Ensembl Genes release 94, GRCh38.p12 assembly). Quality control was performed using the FastQC and RNA-SeQC programs, and samples were included for analysis if they met the following criteria: > 10 million total reads, > 80% of reads mapped to the reference genome, XIST and Y chromosome expression was consistent with reported sex, < 10% of R1 reads in the sense orientation, Pearson correlation ≥ 0.9 with samples in the same library construction batch, and concordant genotype calls between variants called from RNA sequencing reads and DNA genotyping. Sequencing read counts were obtained from the featureCounts function in the Rsubread R package (v1.32.2). Genes were filtered to remove very low expressed genes; we limited our analysis to include only those genes which had an average counts per million (CPM) value > 1 in more than 20 subjects. The trimmed mean of M values (TMM) procedure from the edgeR R package (v3.24.3) was applied to account for the differences in sequencing depth. The gene count data used for this analysis is available in GEO (accession number GSE158699). Counts were transformed to log2 counts per million (CPM) values and quantile-normalized.

### Measurement of plasma protein biomarkers

At Visit 2, plasma samples were assayed for 4,979 proteins in 5,670 COPDGene participants using the SOMAscan Human Plasma 5.0K assay, a multiplex aptamer-based assay (SomaLogic, Boulder, Colorado)[14]. The standardization process included within-plate hybridization, median signal normalization, and plate scaling and calibration of SOMAmers to control for inter-assay variation between analytes and batch differences between plates. Inverse normal transformation was applied to all protein measurements prior to association analysis.

### Cotinine measurements

Plasma metabolite measurements were obtained from plasma for 1,136 subjects at COPDGene Visit 2 using the Metabolon Global Metabolomics Platform (Durham, NC, USA)[15]. Metabolite levels were quantified as raw area counts. Plasma cotinine levels were evaluated, and missing values were assigned a level of half the lowest detected amount of cotinine. Values were log-transformed, and differences in cotinine distribution between groups were evaluated with the Wilcoxon Rank Sum test. COPDGene metabolomic data are available in the <u>Metabolomics Workbench</u> of the NIH Common Fund’s National Metabolomics Data Repository (Study ID ST001443;).

### RNA-seq differential expression and protein association analyses

We used the limma R package (v3.38.3)[16] to test for the associations between RNA transcripts and the vaping/smoking groups, using the former cigarette user group as the reference and adjusting for age, sex, race, and library preparation batch effects. The voom function was used in the RNA-seq analysis. Regression models for protein expression used identical covariates, with the exception that library preparation batch effects were replaced with a variable for clinical center. For both differential gene expression and protein association analyses, we corrected for multiple testing with the Benjamini-Hochberg method and applied a threshold of significance of a false discovery rate (FDR) of 10%[17]. For selected genes and proteins that were associated in only one contrast (vaping or dual only associations), we also conducted association analyses for the contrasts of vapers versus cigarette users, vapers versus dual users, and dual users versus cigarette users. Gene set enrichment analysis were performed using the fast gene set enrichment analysis (FGSEA) R package (v3.12)[18]. Further details are included in the supplementary materials of the online preprint.

### Associations of transcripts and proteins to health outcomes

Biomarkers significantly associated with vaping or dual use were tested for association to prospective health-related outcomes, namely change in FEV1 (absolute volume and percent change from baseline), prospective exacerbations, all-cause and respiratory mortality, and incident CVD. FEV_1_ changes were computed by subtracting Visit 2 from Visit 3 FEV_1_ values and dividing this difference by the number of years between both visits. FEV_1_ changes were calculated as both absolute and relative (as a percentage of the Visit 2 measurement) values. Data points >5 standard deviations from the mean were excluded from analysis.

The FEV1 outcomes were evaluated using multivariable linear regression models, and the time-to-event outcomes were evaluated using Cox-proportional hazards models using Visit 2 as the starting point. Analyses were adjusted for sex, race, and age at baseline (Visit 2). For time-to-event outcomes baseline FEV1 percentage of predicted was included as a covariate. Since the sample sizes of our vaping/smoking groups were small, these association analyses were performed in all available COPDGene study subjects without stratifying by vaping or smoking exposure. The FDR was controlled at 10% using the Benjamini-Hochberg method.

### Prediction modeling for vaping status and prospective health outcomes

Predictive models using only transcript and protein biomarkers were constructed for vaping/smoking groups and prospective health outcomes. For categorical and binary outcomes, linear discriminant analysis (LDA) was used. For change in FEV1 models, elastic net models were constructed. Cross-validation was used to prevent overfitting and estimate predictive performance. Model performance was assessed by cross-validation using the Youden index (sensitivity + specificity - 1), area under the receiver operator characteristic curve (AUROC), and area under the precision recall curve (AUCPR) for binary outcomes. R2 was used to evaluate the performance of change in FEV1 models. Additional details are included in the supplementary materials of the online preprint.

## Results

### Baseline characteristics

A visual overview of the study and data flow is included in Supplemental Figures 1 and 2. There were 51 and 77 subjects reporting current vaping only or dual use, respectively, at the COPDGene second study visit. Differences in baseline characteristics of these groups along with the current and former cigarette user groups are reported in Table 1. The large majority of subjects in the vaping group (80%) reported vaping at least once a day. The dual user group reported lighter vaping on average, with only 42% reporting daily vaping, and their pack-years exposure to combustible tobacco was similar to the current cigarette user group. FEV1 % of predicted values were comparable for all groups. Biochemical confirmation of nicotine exposure via analysis of plasma cotinine levels in 853 subjects demonstrated elevated cotinine levels in the vaping, dual use, and current cigarette user groups relative to the former cigarette user group (Figure 1, Wilcoxon rank sum test p*<*0.005 for all comparisons). A subset of the former cigarette user group had elevated levels of nicotine that could be due to use of nicotine replacement therapy, secondhand exposure to tobacco smoke or e-cigarette aerosols, or unreported smoking or vaping.

**Figure 1.** Plasma cotinine levels by vaping/smoking group. In a subset of subjects, plasma metabolomics data was measured using the Metabolon Global Metabolomics Platform. Violin plots of log-transformed cotinine raw area counts are shown. Cotinine levels are significantly higher in all three groups relative to FS (Wilcoxon rank sum p < 0.005 for all three comparisons). FS = former cigarette users. CS = current cigarette users.

**Table 1.** Baseline characteristics of the vaping and smoking groups

|  | **Current**<br>**cigarette users**<br>**(n = 1,328)** | **Former**<br>**cigarette users**<br>**(n = 2,436)** | **Vapers**<br>**(n = 51)** | **Dual users**<br>**(n = 77)** | **P-**<br>**value** |
| --- | --- | --- | --- | --- | --- |
| Age | 59.6 (9.5) | 68.6 (11.9) | 64.6 (10.2) | 61.6 (9.9) | < 0.001 |
| Sex, % male | 50.5% | 52.0% | 39.2% | 48.1% | 0.3 |
| % Non-Hispanic Whites | 47.1% | 82.4% | 88.2% | 71.4% | < 0.001 |
| BMI | 27.2 (8.4) | 28.7 (7.6) | 29.2 (9.6) | 26.9 (7.1) | < 0.001 |
| Smoking pack-years | 41.1 (25.8) | 38.0 (29.2) | 50.1 (26.3) | 45.0 (26.3) | < 0.001 |
| FEV<sub>1</sub> (L) | 2.2 (1.1) | 2.1 (1.2) | 2.1 (1.2) | 2.1 (1.2) | < 0.001 |
| FEV<sub>1</sub>, % predicted | 81.9 (28.0) | 82.0 (35.6) | 83.2 (32.7) | 76.9 (29.7) | 0.2 |
| FVC (L) | 3.1 (1.4) | 3.0 (1.4) | 3.1 (1.5) | 3.0 (1.5) | 0.02 |
| Use of e-cigarettes: |  |  |  |  |  |
| Less than once a month | NA | NA | 0.0% | 2.6% | 0.3 |
| Less than once a week | NA | NA | 0.0% | 15.6% | 0.003 |
| Some days (1-3 days a week) | NA | NA | 4.0% | 27.3% | < 0.001 |
| Most days | NA | NA | 16.0% | 13.0% | 0.6 |
| Every day | NA | NA | 80.0% | 41.6% | < 0.001 |
| Cartridge size: |  |  |  |  |  |
| 0 mg | NA | NA | 8.2% | 4.0% | 0.3 |
| 6-8 mg | NA | NA | 20.4% | 16.0% | 0.5 |
| 9-12 mg | NA | NA | 14.3% | 9.3% | 0.4 |
| 13-16 mg | NA | NA | 14.3% | 6.7% | 0.2 |
| Do not know | NA | NA | 22.5% | 52.0% | 0.001 |
| Other | NA | NA | 20.4% | 12.0% | 0.2 |
| Number of e-cigarettes used<br>(last 24 hours) | NA | NA | 10 (16) | 3 (10) | < 0.001 |
| Number of cartridges used/week | NA | NA | 2 (6) | 1 (1) | 0.002 |
| E-cigarette brand: |  |  |  |  |  |
| Blu | NA | NA | 4.6% | 17.4% | 0.04 |
| Henley | NA | NA | 2.3% | 1.5% | 0.7 |
| Joye | NA | NA | 0.0% | 2.9% | 0.3 |
| NJOY | NA | NA | 2.3% | 8.7% | 0.2 |
| V2 | NA | NA | 2.3% | 2.9% | 0.8 |
| Other | NA | NA | 90.9% | 71.0% | 0.01 |
| SGRQ Symptom Score | 28.7 (40.4) | 17.9 (39.0) | 15.8 (46.3) | 43.5 (48.5) | < 0.001 |
| Respiratory exacerbation | 14.4% | 17.9% | 21.6% | 23.4% | 0.01 |
| CVD | 18.5% | 25.8% | 15.7% | 18.2% | < 0.001 |

Continuous variables are reported as medians (interquartile ranges). Categorical variables are reported as percentages. Kruskal-Wallis rank sum tests were used for continuous variables. Chi-square tests were used for categorical variables. BMI: Body mass index; CVD: Cardiovascular disease (history of stroke, heart attack, coronary artery disease, coronary artery bypass graft surgery, peripheral artery disease, and/or cardiac angina); Respiratory exacerbation: At least one respiratory exacerbation (acute worsening of respiratory symptoms that required systemic steroids and/or antibiotics) in the previous year; FEV<sub>1</sub>: Forced expiratory volume in 1 second; FEV<sub>1</sub>, % predicted: Observed FEV<sub>1</sub> / Predicted FEV<sub>1</sub> based Hankinson reference equation; FVC: Forced vital capacity; SGRQ: St. George's respiratory questionnaire score. % LAA-950: Percentage of CT low attenuation less than -950 HU at end-inspiration using Thirona software; % Segmental airway wall thickness: Percentage of the wall relative to the total bronchial area for the segmental airways. NA = Not applicable.

### Blood RNA-seq associations with vaping and dual use

A total of 18,303 genes were tested for differential expression analysis of vapers, dual users, and current cigarette users, using former cigarette users as the reference group. The top results from each analysis are shown in Table 2, and complete results are reported in Tables S1-S3. For vapers, three significant associations were identified at an FDR of 10% (Figure 2). Two of these genes (*LINC02470* and *NPHP1)* were expressed at a higher level in vapers than any of the other three groups (p<0.05 for all comparisons, Table S4). The third gene, *GPR15*, was one of the most well-reported genes upregulated by smoking. This gene was also upregulated in vapers, but to a lesser extent than in either cigarette users or dual users (p<0.005 for all comparisons). Gene set enrichment analysis identified 16 significant MSigDB Hallmark pathways with the top three processes being related to inflammation, oxidative phosphorylation, and heme metabolism (Table 3). For all significant gene sets, enrichment was in the direction of decreased expression in vapers compared to former cigarette users. Since smoking has been shown to have mixed effects on inflammation, we compared the pattern of gene set enrichment for the Hallmark Inflammation pathway in vapers, current cigarette users, and dual users, and we observed that smoking and dual use show elements of activation and repression of this gene set; the effect of vaping was more clearly downregulation of this pathway (Supplemental Figure 3).

**Figure 2.** Significantly associated vaping RNA biomarkers. Three RNA biomarkers were significantly associated at 10% FDR in the analysis comparing vapers to former cigarette users. *NPHP1* and *LINC02470* were significantly associated in the vaping but not the smoking or dual use comparison to former cigarette users. *GPR15* is significantly elevated in all three groups relative to former cigarette users, but its expression is relatively lower in vapers relative to dual users and current cigarette users (p<0.005 for all comparisons).

**Table 2.**
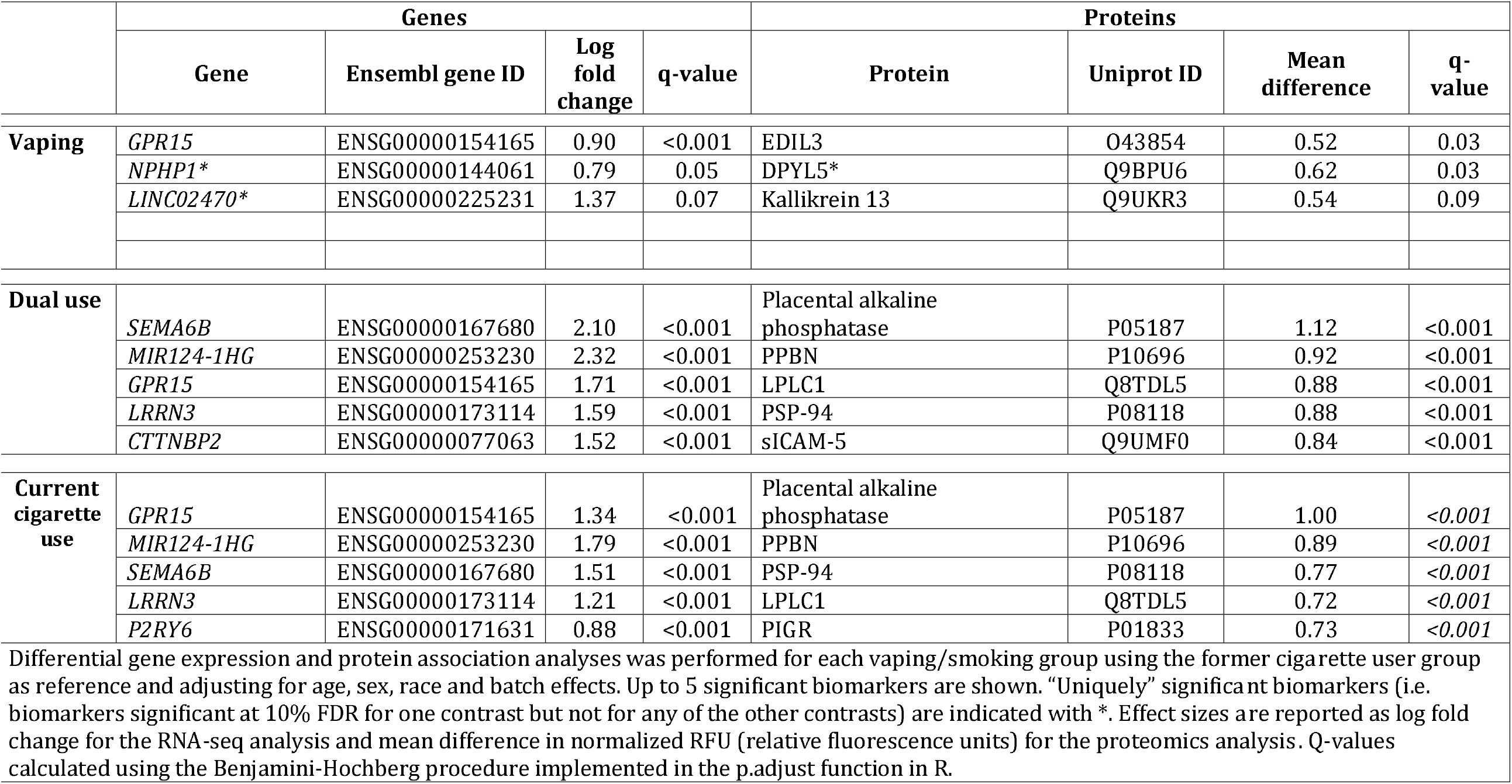
Top 5 differentially associated genes and proteins for vapers, dual users, and current cigarette users.

|  | Genes |  |  |  | Proteins |  |  |  |
| --- | --- | --- | --- | --- | --- | --- | --- | --- |
|  | Gene | Ensembl gene ID | Log fold change | q-value | Protein | Uniprot ID | Mean difference | q-value |
| **Vaping** | *GPR15* | ENSG00000154165 | 0.90 | <0.001 | EDIL3 | O43854 | 0.52 | 0.03 |
|  | *NPHP1\** | ENSG00000144061 | 0.79 | 0.05 | DPYL5* | Q9BPU6 | 0.62 | 0.03 |
|  | *LINC02470\** | ENSG00000225231 | 1.37 | 0.07 | Kallikrein 13 | Q9UKR3 | 0.54 | 0.09 |
| **Dual use** | *SEMA6B* | ENSG00000167680 | 2.10 | <0.001 | Placental alkaline phosphatase | P05187 | 1.12 | <0.001 |
|  | *MIR124-1HG* | ENSG00000253230 | 2.32 | <0.001 | PPBN | P10696 | 0.92 | <0.001 |
|  | *GPR15* | ENSG00000154165 | 1.71 | <0.001 | LPLC1 | Q8TDL5 | 0.88 | <0.001 |
|  | *LRRN3* | ENSG00000173114 | 1.59 | <0.001 | PSP-94 | P08118 | 0.88 | <0.001 |
|  | *CTTNBP2* | ENSG00000077063 | 1.52 | <0.001 | sICAM-5 | Q9UMF0 | 0.84 | <0.001 |
| **Current cigarette use** | *GPR15* | ENSG00000154165 | 1.34 | <0.001 | Placental alkaline phosphatase | P05187 | 1.00 | <0.001 |
|  | *MIR124-1HG* | ENSG00000253230 | 1.79 | <0.001 | PPBN | P10696 | 0.89 | <0.001 |
|  | *SEMA6B* | ENSG00000167680 | 1.51 | <0.001 | PSP-94 | P08118 | 0.77 | <0.001 |
|  | *LRRN3* | ENSG00000173114 | 1.21 | <0.001 | LPLC1 | Q8TDL5 | 0.72 | <0.001 |
|  | *P2RY6* | ENSG00000171631 | 0.88 | <0.001 | PIGR | P01833 | 0.73 | <0.001 |

Differential gene expression and protein association analyses was performed for each vaping/smoking group using the former cigarette user group as reference and adjusting for age, sex, race and batch effects. Up to 5 significant biomarkers are shown. “Uniquely” significant biomarkers (i.e. biomarkers significant at 10% FDR for one contrast but not for any of the other contrasts) are indicated with \*. Effect sizes are reported as log fold change for the RNA-seq analysis and mean difference in normalized RFU (relative fluorescence units) for the proteomics analysis. Q-values calculated using the Benjamini-Hochberg procedure implemented in the p.adjust function in R.

**Table 3.** Top 5 significant (adjusted P-values < 0.1) MSigDB Hallmark pathways from differential gene expression analyses for the vaping and smoking groups.

|  | **MSigDB Hallmark pathway** | **Pathway ID** | **Number of genes** | **NES** | **Adjusted P-value** |
| --- | --- | --- | --- | --- | --- |
| **Vaping** | Inflammatory Response | M5932 | 163 | -4.10 | $6.3 \times 10^{-10}$ |
| | Oxidative Phosphorylation | M5936 | 200 | -4.01 | $2.8 \times 10^{-9}$ |
| | Heme Metabolism | M5945 | 195 | -3.82 | $2.7 \times 10^{-8}$ |
| | TNF-α Signaling via NF-kB | M5890 | 179 | -3.85 | $3.7 \times 10^{-8}$ |
| | Complement | M5921 | 171 | -3.50 | $4.0 \times 10^{-7}$ |
| **Dual Use** | Heme Metabolism | M5945 | 195 | -5.53 | $3.9 \times 10^{-17}$ |
| | Oxidative Phosphorylation | M5936 | 200 | -3.70 | $1.7 \times 10^{-7}$ |
| | E2F Targets | M5925 | 199 | -3.69 | $2.0 \times 10^{-7}$ |
| | Interferon gamma response | M5913 | 194 | -2.90 | $1.4 \times 10^{-4}$ |
| **Current cigarette use** | Heme Metabolism | M5945 | 195 | -8.68 | $1.2 \times 10^{-48}$ |
| | MYC Targets V1 | M5926 | 200 | 3.33 | $6.1 \times 10^{-6}$ |
| | Estrogen Response Late | M5907 | 145 | -2.89 | $2.2 \times 10^{-4}$ |
| | Xenobiotic Metabolism | M5934 | 151 | -2.86 | $3.2 \times 10^{-4}$ |
| | Epithelial mesenchymal transition | M5930 | 129 | -2.86 | $3.8 \times 10^{-4}$ |

Gene set enrichment analysis (GSEA) was performed using the results from the differential expression analysis comparing vapers, dual users, or current cigarette users to former cigarette users. Genes were ranked for enrichment analysis by log fold change. The top 5 results from the MSigDB Hallmarks gene sets are shown ordered by p-value. The “number of genes” column is the number of genes associated with the GSEA pathway that could be mapped to the differential expression results. Abbreviations: NES = normalized enrichment score. UV= ultraviolet light.

For dual users, 90 differentially expressed genes were identified, and 17 of these genes were significant only in the dual user analysis. All 17 genes were also significantly differentially expressed when comparing dual users to current cigarette users, suggesting that these genes have a distinctive response to dual use (p<0.005 for all comparisons, see Table S5). Gene set enrichment analysis identified four significant Hallmark pathways related to heme metabolism, oxidative phosphorylation, E2F signaling, and interferon gamma response (Table 3). Complete results for the vaping, dual use, and smoking analyses including MSigDB Immunologic Signature pathways are presented in Supplemental Tables S6-S8.

### Proteomic biomarker associations to vaping and dual use

A total of 4,979 SOMAscan aptamers mapping to 4,720 unique proteins were tested for association to vaping and smoking groups. The top significantly associated proteins are shown in Table 2 (see Table S9 for complete results), and violin plots for selected top protein biomarkers associated with vaping or dual use are shown in Figure 3. Three protein biomarkers (EDIL3, DPYL5, and KLK13) were significantly associated with vaping status at a 10% FDR, and DPYL5 expression was found to be higher in vapers compared to each of the other groups (p<0.05 for all comparisons, Table S10). One hundred protein biomarkers were associated with dual use, with four (CXA8, Keratin 19, CSKP, and PXDC1) being significantly associated only with dual use. All four of these proteins also showed nominal association (p<0.05) in an analysis comparing dual users to current cigarette users, indicating a distinct response to dual use (Table S11).

**Figure 3.** Top protein biomarkers significantly associated with vaping or dual use. The distribution of the top significantly associated protein biomarkers in the analysis of vapers or dual users versus former cigarette users. DPYL5 is significantly elevated in vapers relative to all other vaping/smoking groups (p<0.005). KLK13 is significantly associated with vapers, dual users, and current cigarette users when compared to former cigarette users. s-ICAM5 and Secretoglobin family member 3A member 1 were significantly associated with dual users and current cigarette users but not vapers when compared to former cigarette users.

### Comparison of biomarker associations to vaping, smoking, and dual use

To compare the similarity of the transcriptomic and proteomic responses to each exposure, we constructed pairwise plots of the effect sizes of all significantly associated biomarkers (Figure 4). There was a strong positive correlation between the biomarker responses to smoking and dual use (Pearson’s r = 0.76 and 0.88 for RNA and protein, respectively), whereas the vaping biomarker profile had low correlation to the profiles for both dual use and smoking (r between 0.23 and 0.44). Certain biomarkers (LINC02470 and DPYL5 protein) had particularly strong and unique associations with vaping. When we compared effect sizes between RNA and proteomic biomarkers associated with vaping and smoking, the correlation was low (r=0.11) as might be expected given various potential sources of origin for circulating blood proteins.

**Figure 4.** Correlation of biomarker responses between vapers, dual users, and current cigarette users. Comparison of the biomarker association profiles demonstrates that while the dual user and current cigarette user profiles are very similar, the vaping biomarker profile is more distinct. For the RNA and protein biomarker analysis of vaping/smoking groups versus former cigarette users, the effect sizes for each pair of comparisons were plotted and the Pearson correlation of effect sizes was calculated. In each pairwise comparison, only biomarkers significant in at least one of analyses were analyzed. In row A, vaping-associated RNA biomarker effects are compared to current cigarette user effects (left), dual user effects (middle), and then dual user and current cigarette user effects are compared (right). In row B, the corresponding protein biomarker effects are compared.

### Classification accuracy for vaping/smoking behavior using biomarker panels

To determine how well RNA and protein biomarkers could classify subjects according to their vaping/smoking behavior, we used a nested cross-validation approach to provide an unbiased estimate of the classification performance achievable by multi-class prediction models using linear discriminant analysis (LDA). The performance of this model is shown in Table 4, where for each group the task was to distinguish that group from all other groups combined. The model achieved good discrimination for current cigarette users (Youden index (YI) = 0.81) and former cigarette users (YI = 0.80) and had statistically significant but less accurate performance for vapers (YI = 0.53) and dual users (YI = 0.55). More detailed metrics for each pairwise contrast (Table S12) demonstrates that the models do better in discriminating vapers from current cigarette users than former cigarette users (YI = 0.73 and 0.50, respectively), whereas for dual users, model discrimination was better for former cigarette users (YI = 0.82) than current cigarette users (YI = 0.34), providing further evidence of the similarity between dual use and current smoking biomarker profiles.

**Table 4.** Multi-class prediction of vaping and smoking groups using multi-omic biomarkers

|  | Integrated RNA-seq and proteomic data |  |  |  |
| --- | --- | --- | --- | --- |
|  | Current cigarette users | Former cigarette users | Vapers | Dual users |
| AUROC | 0.94 | 0.94 | 0.71 | 0.71 |
| AUPRC | 0.85 | 0.97 | 0.06 | 0.04 |
| Youden index | 0.81 | 0.80 | 0.53 | 0.55 |
| Linear discrimination analysis models were used to classify vaping and smoking groups. For each group, LDA models are constructed to distinguish that group from the other three using 10-fold nested cross validation (CV) to estimate performance. Abbreviations: AUROC = Area under the receiving operating characteristic curve; AUPRC = Area under the precision-recall curve. The Youden Index (YI) metric was used to summarize the model performance at the optimal cutoff on the area under the receiver operating curve. |  |  |  |  |

### Vaping and dual use biomarkers and longitudinal outcomes

To determine whether biomarkers of vaping or dual use were associated with prospective health-related outcomes, we tested the 92 RNA transcripts and 101 protein biomarkers associated with vaping and/or dual use for association to multiple health outcomes in all COPDGene subjects. Twenty-one transcripts and 88 protein biomarkers were associated with one or more of the studied outcomes at a 10% FDR level. Of the vaping-associated biomarkers, two (KLK13 and DPYL5) were associated with loss of FEV1, and KLK13 was also associated with mortality. Boxplots for the top four vaping or dual use proteins associated with all-cause mortality are shown in Figure 5, and all significant biomarker associations are shown in Tables S13-S16.

**Figure 5.** Top protein vaping and dual use biomarkers significantly associated with overall mortality. The dual use-associated proteins Growth hormone receptor, MIC-1, and HE4 were significantly associated with overall mortality. KLK-13 was the only vaping associated biomarker that was also associated mortality. All associations were significant at a false discovery rate of 10%.

We then determined whether the effect of vaping or dual use on each protein biomarker was consistent with higher or lower risk for each adverse health outcome, and we observed that the direction of the exposure effect was overwhelmingly towards increased risk for adverse health events (Figure 6). Out of 246 total significant associations to health outcomes, 224 (91%) showed concordance between the effect of vaping or dual use and the direction of association to increased risk. Both of the vaping-associated biomarkers (KLK13 and DPYL5) showed concordant effects for higher health risk.

**Figure 6.** Concordance analysis of biomarker associations to vaping/dual use and adverse health outcomes. For protein biomarkers that were significantly associated with at least one health outcome and vaping (n=2) or dual use (n=86), we plotted the effect of the exposure (vaping or dual use) against the effect with respect to the health outcome. Since some biomarkers were significantly associated with multiple health outcomes, the total number of analyzed associations was 246. The top panel shows the relationship between exposure effect and change FEV1 (ml/yr) where each of the 107 associations was concordant for loss of FEV1. Change in FEV1 is calculated as visit 3 value - visit 2 value, meaning negative values indicate loss of lung function. The bottom panel shows the relationship between exposure effect and hazard ratio for all cause mortality, respiratory mortality, exacerbations, or incident cardiovascular disease. 117 of 139 associations were concordant for higher health risk. Exposure effect is the beta coefficient from the dual use versus former cigarette users analysis, except for DPYL5 and KLK13 which is from the vapers versus former cigarette users analysis.

To estimate the accuracy of multi-marker panels of vaping and dual use biomarkers for predicting prospective health outcomes, we constructed predictive models for FEV1 decline, all-cause mortality, COPD exacerbations, and incident CVD using only the vaping or dual use-associated RNA and protein biomarkers. Models using the seven vaping-associated biomarkers alone gave non-informative predictions, but models using the 190 dual use-associated biomarkers gave informative predictions with moderate accuracy for FEV1 decline, all-cause mortality and respiratory exacerbations (Table 5).

**Table 5.** Performance metrics of prediction models for prospective health outcomes.

| Biomarker Panel | Outcome | Sample size (events) | AUROC | AUPR | Youden Index | R2 |
| --- | --- | --- | --- | --- | --- | --- |
| Dual Use Biomarkers | Δ FEV1 (ml/yr) | 642 | NA | NA | NA | 0.08 |
|  | Δ FEV1 (% of baseline) | 642 | NA | NA | NA | 0.17 |
|  | Overall mortality | 2388 (246) | 0.68 | 0.38 | 0.36 | NA |
|  | Exacerbation >= 1 | 2231 (811) | 0.63 | 0.51 | 0.25 | NA |
|  | Incident CVD | 1700 (183) | 0.68 | 0.34 | 0.36 | NA |
| Prediction models for the indicated outcomes were constructed using elastic net for change in FEV1 models and linear discriminant analysis for binary outcomes. Δ FEV1 (ml/yr) = (FEV1 at visit 3 - FEV1 at visit 2) / years between visits. Δ FEV1 (% of baseline) = Δ FEV1 (ml/yr) / FEV1 at visit 2. Exacerbation = 1 if subjects reported one or more exacerbation over the follow-up period after visit 2 and 0 otherwise. Incident CVD = 1 if subject reported new diagnosis of CVD after visit 2, and subjects with existing CVD at visit 2 were excluded. AUROC = area under the receiver operator characteristic. AUPR = area under the precision-recall curve. R2 = r-squared. |  |  |  |  |  |  |

### Replication of previously reported vaping biomarker associations

A recently published review of vaping biomarkers listed 14 inflammatory and matrix degradation protein biomarkers that had been previously associated with vaping[19]. We examined the results from these 14 biomarkers at the RNA and protein level for association to vaping or dual use, and we observed associations at nominal significance (p<0.05) for four biomarkers (*IL1B*, ICAM1 [both RNA and protein], IL-6, and MMP9, see Table S17).

## Discussion

Identifying validated biomarkers of vaping and dual use is an important tool both for guiding ENDS users as to the risks of these devices and effective regulation of ENDS. This study of established adult cigarette users identified significant associations of six and 190 blood molecular markers with vaping and dual use, respectively. Because of the comprehensive nature of the ‘omics measurements, we were able to compare overall molecular profiles of vaping, dual use, and smoking. These analyses found that the vaping profile was substantially different from current smoking, whereas dual use and smoking were similar but not identical. Biomarkers associated with vaping and dual use were also associated with multiple cardiopulmonary health outcomes, and reasonably predictive accuracy for some of these outcomes could be achieved for models using dual use biomarkers.

A previous metabolomic biomarker study by Goniewicz *et al*. demonstrated that vaping is associated with metabolomic changes, albeit at generally lower levels compared to current cigarette users or dual users[20]. A recent study of vapers versus non-cigarette users identified numerous changes in plasma biomarkers associated with inflammation, oxidative stress, and extracellular matrix degradation[19]. A recent meta-analysis of biomarkers of ENDS versus cigarettes reported a more favorable biomarker profile of ENDS use, though the last author of the study has been reported to have ties to the tobacco industry[21,22]. Another systematic review also reported reduced levels of some biomarkers in ENDS users relative to current cigarette users, but this is not consistent for all biomarkers (Hiler *et al*., 2021). By examining the correlation in biomarker profiles for vaping, dual use, and smoking, we observed that the vaping profile is largely distinct from both the dual use and smoking profile which are in turn highly correlated to each other. We did however observe multiple examples of shared RNA and protein biomarkers between smoking, vaping, and dual use, such as *GPR15*, a regulator of inflammation and T-cell trafficking. We also provided independent validation for previously reported vaping or dual use associations with *IL1B*, ICAM1, IL-6, and MMP9, all of which are known to influence inflammation (IL-1B can induce MMP-9 expression and activity).

Both vaping and dual use are associated with diverse effects on biological pathways related to aspects of inflammation, oxidative phosphorylation, and coagulation-related pathways. Previous studies have also shown that long non-coding RNAs (lncRNAs) are associated with vaping[23,24], and in our work we identified LINC02470 as a novel vaping biomarker. LINC02470 has previously been reported to modulate Wnt-beta catenin signaling via exosomal secretion from bladder cancer cells[25]. Because Tang *et al*. previously demonstrated bladder urothelial hyperplasia in mice exposed daily to e-cigarette aerosols, the finding of LINC02470 in the circulation of human ENDS users is highly concerning. We also identified DPYL5 as being uniquely upregulated in vaping subjects. DPYL5 is primarily expressed in brain tissue and glial cells, though it is also expressed at detectable levels in the gastrointestinal (GI) tract. DPYL5 is potentially an important vaping biomarker, because it was also predictive of future loss of lung function. To our knowledge, this is the first report of DPYL5 as a specific and clinically relevant biomarker of vaping, a finding that requires further independent validation.

Many of the biomarkers altered by vaping and dual use are also associated with risk of adverse health events such as loss of lung function, respiratory exacerbations, and mortality. For the overwhelming majority of biomarkers, the direction of effect of vaping and dual use was consistent with increased health risk. This extends numerous prior observations linking vaping to a variety of pulmonary symptoms and alterations of cardiovascular physiology[26], providing an important link to longer-term health outcomes and suggesting that some of these biomarkers may be useful in assessing health risks of specific vaping devices and e-liquid formulations. For dual use, a multi-marker panel of associated biomarkers achieved modest predictive accuracy for some important outcomes. For vapers, the biomarker profile was more subtle, and with our current sample size we did not identify a sufficient number of biomarkers to enable accurate multi-marker prediction of health outcomes.

The strengths of this study are the high-throughput approach to biomarker discovery, the characterization of associations to prospective health events, and the use of machine learning to assess the predictive performance of multimarker panels. Limitations include a modest sample size of vapers and dual users and the lack of independent replication due to lack of available cohorts with similar biomarker data and exposure characterization. Information regarding specific vaping devices and fluids was limited and reflects the challenges posed by the rapid evolution of vaping devices over the study period. Our findings cannot be assumed to generalize to newer-generation vaping devices which will require dedicated study, and our findings in adults do not necessarily apply to other important groups such as adolescents.

In conclusion, this study identified over one-hundred transcriptomic and proteomic biomarkers associated with vaping and dual use, and many of these biomarkers are also associated with prospective cardiopulmonary health outcomes. Prediction of health outcomes using multimarker panels can provide informative prediction with room for improvement, and validation in independent cohorts will increase confidence in individual biomarkers and multi-marker models. Larger-scale studies with greater power would be likely to identify additional biomarkers that could further improve predictive model performance.

## Data Availability

All data produced are available online at dbGaP

https://www.ncbi.nlm.nih.gov/projects/gap/cgi-bin/study.cgi?study_id=phs000179.v6.p2

## Data availability

Supplemental materials are included in the medRxiv online preprint (doi: https://doi.org/10.1101/2022.09.19.22280093).

### Underlying data

NCBI dbGAP: https://www.ncbi.nlm.nih.gov/projects/gap/cgi-bin/study.cgi?study_id=phs000179.v6.p2

## References

1 Gentzke AS, Wang TW, Jamal A, et al. Tobacco Product Use Among Middle and High School Students - United States, 2020. MMWR Morb Mortal Wkly Rep 2020;69:1881–8.

2 Mayer M, Reyes-Guzman C, Grana R, et al. Demographic Characteristics, Cigarette Smoking, and e-Cigarette Use Among US Adults. JAMA Netw Open 2020;3:e2020694.

3 Li D, Sundar IK, McIntosh S, et al. Association of smoking and electronic cigarette use with wheezing and related respiratory symptoms in adults: cross-sectional results from the Population Assessment of Tobacco and Health (PATH) study, wave 2. Tobacco Control. 2019;:tobaccocontrol – 2018. doi:10.1136/tobaccocontrol-2018-054694

4 Yao T, Max W, Sung H-Y, et al. Relationship between spending on electronic cigarettes, 30-day use, and disease symptoms among current adult cigarette smokers in the U.S. PLoS One 2017;12:e0187399.

5 St Helen G, Eaton DL. Public Health Consequences of e-Cigarette Use. JAMA Intern Med 2018;178:984–6.

6 Smith ML, Gotway MB, Crotty Alexander LE, et al. Vaping-related lung injury. Virchows Arch 2021;478:81–8.

7 Cherian SV, Kumar A, Estrada-Y-Martin Rm. E-Cigarette or Vaping Product-Associated Lung Injury: A Review. Am J Med 2020;133:657–63.

8 Hammond D, Reid JL, Rynard VL, et al. Prevalence of vaping and smoking among adolescents in Canada, England, and the United States: repeat national cross sectional surveys. BMJ 2019;365:l2219.

9 Miech R, Johnston L, O’Malley PM, et al. Trends in Adolescent Vaping, 2017–2019. N Engl J Med 2019;381:1490–1.

10 Miech R, Johnston L, O’Malley PM, et al. Adolescent vaping and nicotine use in 2017-2018 - U.s. national estimates. N Engl J Med 2019;380:192–3.

11 Aronson JK. Research priorities in biomarkers and surrogate end-points. Br J Clin Pharmacol 2012;73:900–7.

12 FDA-NIH Biomarker Working Group. BEST (Biomarkers, EndpointS, and Other Tools) Resource. Food and Drug Administration (US) 2016.

13 Regan EA, Hokanson JE, Murphy JR, et al. Genetic epidemiology of COPD (COPDGene) study design. COPD 2010;7:32–43.

14 Gold L, Ayers D, Bertino J, et al. Aptamer-based multiplexed proteomic technology for biomarker discovery. PLoS One 2010;5:e15004.

15 Gillenwater LA, Kechris KJ, Pratte KA, et al. Metabolomic Profiling Reveals Sex Specific Associations with Chronic Obstructive Pulmonary Disease and Emphysema. Metabolites 2021;11. doi:10.3390/metabo11030161

16 Ritchie ME, Phipson B, Wu D, et al. limma powers differential expression analyses for RNA-sequencing and microarray studies. Nucleic Acids Res 2015;43:e47.

17 Benjamini Y, Hochberg Y. Controlling the false discovery rate: A practical and powerful approach to multiple testing. J R Stat Soc 1995;57:289–300.

18 Gennady Korotkevich, Vladimir Sukhov, Nikolay Budin, Boris Shpak, Maxim N. Artyomov, Alexey Sergushichev. An algorithm for fast preranked gene set enrichment analysis using cumulative statistic calculation. biorxiv. 2021. doi:10.1101/060012

19 Singh KP, Lawyer G, Muthumalage T, et al. Systemic biomarkers in electronic cigarette users: implications for noninvasive assessment of vaping-associated pulmonary injuries. ERJ Open Res 2019;5. doi:10.1183/23120541.00182-2019

20 Goniewicz ML, Smith DM, Edwards KC, et al. Comparison of Nicotine and Toxicant Exposure in Users of Electronic Cigarettes and Combustible Cigarettes. JAMA Netw Open 2018;1:e185937.

21 Akiyama Y, Sherwood N. Systematic review of biomarker findings from clinical studies of electronic cigarettes and heated tobacco products. Toxicology Reports. 2021;8:282–94. doi:10.1016/j.toxrep.2021.01.014

22 Neil Sherwood. TobaccoTactics. 2020.https://tobaccotactics.org/wiki/neil-sherwood/ (accessed 26 Jun 2022).

23 Tommasi S, Caliri AW, Caceres A, et al. Deregulation of Biologically Significant Genes and Associated Molecular Pathways in the Oral Epithelium of Electronic Cigarette Users. Int J Mol Sci 2019;20. doi:10.3390/ijms20030738

24 Kaur G, Singh K, Maremanda KP, et al. Differential plasma exosomal long non-coding RNAs expression profiles and their emerging role in E-cigarette users, cigarette, waterpipe, and dual smokers. PLoS One 2020;15:e0243065.

25 Huang C-S, Ho J-Y, Chiang J-H, et al. Exosome-Derived LINC00960 and LINC02470 Promote the Epithelial-Mesenchymal Transition and Aggressiveness of Bladder Cancer Cells. Cells 2020;9. doi:10.3390/cells9061419

26 Bozier J, Chivers EK, Chapman DG, et al. The Evolving Landscape of e-Cigarettes: A Systematic Review of Recent Evidence. Chest 2020;157:1362–90.

