## Supplemental Materials for "Blood RNA and protein biomarkers are associated with vaping and dual use, and prospective health outcomes"

^1^Channing Division of Network Medicine, Brigham and Women’s Hospital, Harvard Medical School, Boston, MA; ^2^Department of Biostatistics, National Jewish Health, Denver, CO; ^3^Division of Pulmonary and Critical Care Medicine, Brigham and Women’s Hospital, Harvard Medical School, Boston, MA; ^4^Division of Pulmonary, Critical Care and Sleep Medicine, National Jewish Health, Denver, CO; ^5^Division of Pulmonary, Critical Care, Physiology and Sleep Medicine, University of California San Diego (UCSD), La Jolla, CA; ^6^Pulmonary Critical Care Section, Veterans Affairs (VA) San Diego Healthcare System, La Jolla, CA; ^7^Division of General Medicine and Primary Care, Brigham and Women’s Hospital, Harvard Medical School, Boston, MA

* Equal contribution

**Corresponding Author:** Peter J. Castaldi, MD, MSc, Channing Division of Network Medicine, Brigham and Women’s Hospital, 181 Longwood Avenue, Boston, MA 02115, Email: [](about:blank)

**Table of Contents**

1) **Supplemental Methods**
2) **Supplemental Tables**
   a) **ST 1. Whole blood RNA differential expression analysis for vapers versus former cigarette users.** (see file Table_S1toS3_RNAseqResults.xlsx)
   b) **ST 2. Whole blood RNA differential expression analysis for dual users versus former cigarette users.** (see file Table_S1toS3_RNAseqResults.xlsx)
   c) **ST 3. Whole blood RNA differential expression analysis for current versus former cigarette users.** (see file Table_S1toS3_RNAseqResults.xlsx)
   d) **S4. Differential expression analysis for all vaper group contrasts for genes significantly associated only to vaping in the primary analyses.** (see file Table_S4toS5_VapingandDualUse_RNAcontrasts.xlsx)
   e) **S5. Differential expression analysis for all dual user group contrasts for genes significantly associated only to dual use in the primary analyses.** (see file Table_S4toS5_VapingandDualUse_RNAcontrasts.xlsx)
   f) **ST6. Gene set enrichment analysis for the vapers vs former cigarette users from RNA-seq differential expression analysis.** (see file Table_S6toS8_RNA_GSEA.xlsx)
   g) **ST7. Gene set enrichment analysis for the dual users vs former cigarette users from RNA-seq differential expression analysis.** (see file Table_S6toS8_RNA_GSEA.xlsx)
   h) **ST8. Gene set enrichment analysis for the current vs former cigarette users from RNA-seq differential expression analysis.** (see file Table_S6toS8_RNA_GSEA.xlsx)
   i) **S9. Protein differential expression results by vaping/smoking group.** (see file TableS9_protein_DE_results.xlsx)
   j) **S10. Differential expression analysis for all vaping contrasts for proteins significantly associated only to vaping in the primary analyses.** (see file Table_S11toS12_VapingandDualUse_Proteincontrasts.xlsx)
   k) **S11. Differential expression analysis for all dual use contrasts for proteins significantly associated only to dual in the primary analyses.** (see file Table_S11toS12_VapingandDualUse_Proteincontrasts.xlsx)
   l) **S12. Pairwise comparisons of vaping and smoking groups using both RNA-seq and proteomic biomarkers.** (see file Table_S12_LDA_Vaping_AllPairwise_noSE.docx)
   m) **S13. RNA Biomarker Associations to 5-year Change in FEV1.** (see file TableS13to16_OutcomeAssociations.xlsx)
   n) **S14. Protein Biomarker Associations to 5-year Change in FEV1.** (see file TableS13to16_OutcomeAssociations.xlsx)
   o) **S15. RNA Biomarker Associations to Time to Event Outcomes.** (see file TableS13to16_OutcomeAssociations.xlsx)
   p) **S16. Protein Biomarker Associations to Time to Event Outcomes.** (see file TableS13to16_OutcomeAssociations.xlsx)
   q) **S17. Previously reported RNA and protein biomarker associations to vaping or dual use.** (see file TableS17_PriorBiomarkers.xlsx)
3) **Supplemental Figures**
   a) **SF1. Study overview.**
   b) **SF2. Flow diagram for study subjects.**
   c) **SF3. GSEA enrichment plots for Hallmark Inflammation Pathway**

**Supplemental Methods:**

***Definitions of exacerbations, incident comorbidities, and mortality assessment***

Respiratory exacerbations were defined as the acute worsening of respiratory symptoms that required systemic antibiotics and/or steroids as determined by self-report. Respiratory disease-related health impairments and quality of life were assessed using the St George’s Respiratory Questionnaire (SGRQ). Incident self-reported cardiovascular disease (CVD) was defined as a composite endpoint of stroke, heart attack, coronary artery disease, coronary artery bypass graft surgery, peripheral artery disease, or cardiac angina. All-cause mortality was determined through longitudinal follow-up and by referencing the social security death index. A systematic adjudication process based on methods outlined in the Towards a Revolution in COPD Health (TORCH) trial was used to categorize cause-specific mortality[[15]](https://paperpile.com/c/BPlaru/rPfiE).

***RNA-sequencing***

The extraction protocol for total blood RNA was performed either manually or with the Qiagen QIAcube extraction robot according to the company’s standard operating procedure. RNA samples with an RNA integrity number (RIN) > 6 and a concentration of ≥ 25 μg/μl were sequenced. Library preparation was performed with the Illumina TruSeq Stranded Total RNA with Ribo-Zero Globin kit (Illumina, Inc., San Diego, CA). Samples were sequenced to an average depth of 20 million 75 base pairs (bp) paired end reads on Ilumina HiSeq 2500 sequencers. Reads were aligned to the GRCh38 genome using the STAR (version 2.5.2b) aligner(1). Gene annotations and transcript GTF were downloaded from the Biomart Ensembl database (Ensembl Genes release 94, GRCh38.p12 assembly). Quality control was performed using the FastQC and RNA-SeQC programs, and samples were included for analysis if they met the following criteria: > 10 million total reads, > 80% of reads mapped to the reference genome, XIST and Y chromosome expression was consistent with reported sex, < 10% of R1 reads in the sense orientation, Pearson correlation ≥ 0.9 with samples in the same library construction batch, and concordant genotype calls between variants called from RNA sequencing reads and DNA genotyping. Sequencing read counts were obtained from the featureCounts function in the Rsubread R package (v1.32.2). Genes were filtered to remove very low expressed genes; we limited our analysis to include only those genes which had an average counts per million (CPM) value > 1 in more than 20 subjects. The trimmed mean of M values (TMM) procedure from the edgeR R package (v3.24.3) was applied to account for the differences in sequencing depth(2).

***Gene set enrichment analysis***

Enrichment analyses for RNA transcripts were performed using the preranked gene set enrichment analysis method reimplemented in the FGSEA R package (v1.16.0)(3), using as gene sets the Hallmark (h.all.v7.5.1) and Immunologic signature (c7.immunesigdb.v7.5.1) pathways from the Molecular Signatures Database. Genes from each differential expression analysis were ranked by their t statistics and the enrichment scores were computed from unweighted ranks by setting the gseaParam argument to 0. The minimal and maximal size of the tested pathways were 15 and 500, respectively. The number of permutations was set to 10,000. The threshold for significant pathways from the FGSEA analyses was an adjusted (Benjamini-Hochberg method) P-value < 0.1.

***Prediction modeling for vaping status and prospective health outcomes***

Predictive models using only transcript and protein biomarkers were constructed for vaping/smoking groups and prospective health outcomes. For the vaping/smoking group prediction models we used linear discriminant analysis (LDA) machine learning for the multiclass classification into the four vaping/smoking groups using nested cross-validation. To avoid bias induced by feature selection, all genes were considered and feature selection was performed within each fold of the inner CV loop. For prediction of prospective binary outcomes, we predicted occurrence of the outcome (binary 1/0) during the follow-up period using LDA using only the transcripts or proteins that were significantly associated with vaping or dual use. For prediction of change in FEV1, the same biomarker predictors were used in an elastic net model. Model performance was assessed by cross-validation using Youden index, area under the receiver operator characteristic (AUROC), and area under the precision recall curve (AUCPR) for binary outcomes. R2 was used for change in FEV1 models. Prediction models were not made for respiratory mortality due to the low number of observed events.

**Supplemental Figures**

**SF 1. Study overview.** The objectives of the present study were to identify blood transcriptomic and proteomic biomarkers of vaping, relate them to prospective health outcomes, and investigate their ability to accurately distinguish vaping and smoking status and to predict prospective health events.

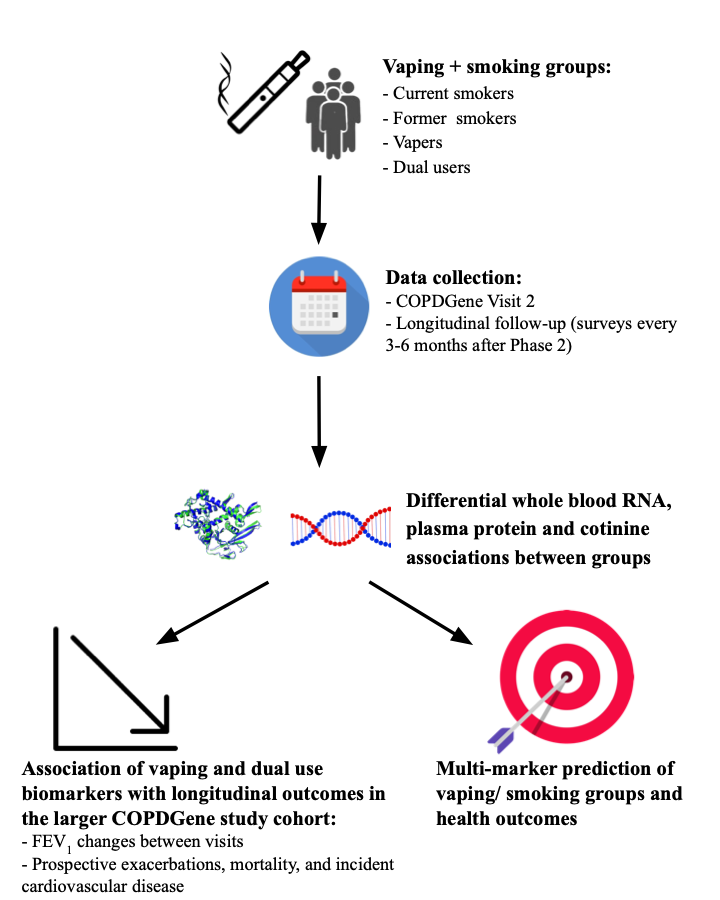

**SF2. Study flow diagram.**

**SF3. GSEA enrichment plots for Hallmark Inflammation Pathway.** Gene set enrichment plots for the Hallmark Inflammation pathway in the RNA-seq analyses comparing vapers (left panel), current cigarette users (middle panel), and dual users (right panel) to former cigarette users as the common control group.

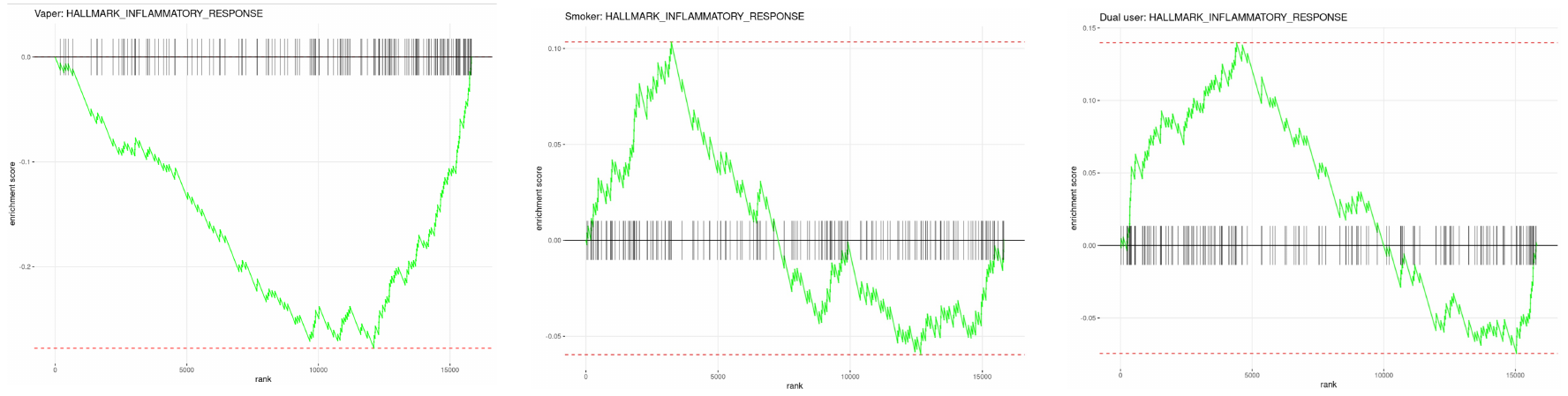

**References**

1.	Dobin A, Davis CA, Schlesinger F, Drenkow J, Zaleski C, Jha S, Batut P, Chaisson M, Gingeras TR. STAR: ultrafast universal RNA-seq aligner. *Bioinformatics* 2013;29:15–21.

2.	Robinson MD, McCarthy DJ, Smyth GK. edgeR: a Bioconductor package for differential expression analysis of digital gene expression data. *Bioinformatics* 2010;26:139–140.

3.	Gennady Korotkevich, Vladimir Sukhov, Nikolay Budin, Boris Shpak, Maxim N. Artyomov, Alexey Sergushichev. An algorithm for fast preranked gene set enrichment analysis using cumulative statistic calculation. biorxiv. 2021. doi:10.1101/060012
