## Supplemental Table 12 for "Blood RNA and protein biomarkers are associated with vaping and dual use, and prospective health outcomes"

**Table S12.** **Pairwise comparisons of vaping and smoking groups using both RNA-seq and proteomic biomarkers.**

|  | | Integrated RNA-seq and proteomic data | | |
| --- | --- | --- | --- | --- |
|  |  | Vapers | Current smokers | Dual users |
| AUROC | Current smokers | 0.860 | - | - |
|  | Dual users | 0.751 | 0.518 | - |
|  | Former smokers | 0.681 | 0.954 | 0.908 |
| AUCPR | Current smokers | 0.348 | - | - |
|  | Dual users | 0.792 | 0.065 | - |
|  | Former smokers | 0.072 | 0.887 | 0.272 |
| Youden’s index | Current smokers | 0.728 | - | - |
|  | Dual users | 0.546 | 0.337 | - |
|  | Former smokers | 0.502 | 0.838 | 0.822 |
| Linear discrimination analysis using ten-fold nested cross validation prediction statistics for pairwise comparisons between the four vaping and smoking groups. Any current smoking behavior included both current smokers and dual users (any non-current smoking behavior included vapers and former smokers), while any vaping behavior included vapers and dual users (any non-vaping behavior included current smokers and former smokers). Means (standard errors) are reported. Abbreviations: AUROC = Area under the receiver operator characteristic; AUCPR = Area under the precision-recall curve. | | | | |
